# Estimating geographic access to health care services for children unvaccinated against diphtheria, tetanus, pertussis across low- and middle-income countries in 2021

**DOI:** 10.64898/2025.12.18.25342640

**Authors:** Natalia Tejedor-Garavito, Maksym Bondarenko, Adelle Wigley, Aubrey Steingraber, C. Edson Utazi, Danielle Boyda, Josh Lorin, Dan Hogan, Jessica Espey, Andrew J. Tatem

## Abstract

Achieving routine immunization for all children remains a major challenge in many low- and middle-income countries. In 2021, 18.1 million children globally missed their first dose of the diphtheria-tetanus-pertussis vaccine (DTP1), highlighting gaps in immunization and healthcare services. The Global Immunization Agenda 2030 aims for 90% coverage of essential childhood and adolescent vaccines and a 50% reduction in zero-dose children by 2030. However, national level data often obscures subnational variations, hampering the identification of unvaccinated children. Geographic access to healthcare is a key factor in vaccine uptake, but global comparative studies at the subnational level are rare, limiting our understanding of local vaccination behaviours and dynamics.

This study estimates and maps the number and distribution of zero-dose children and their geographic access to healthcare services across 99 low- and middle-income countries. Using geospatial modelling approaches, we assessed geographic access to the nearest facility through two scenarios: walking and taking motorised transport. We quantified spatial inequalities of immunization at 1km spatial resolution and compared the patterns with healthcare access.

Results show substantial variations, both between and within countries. Among the estimated 15.7 million zero-dose children across the studied countries, 39% lived more than one hour’s walking from a health facility, with Afghanistan (84%), Papua New Guinea (83%), Sudan (81%), and Cambodia (78%) showing the highest proportions. When figures were disaggregated to the district level, 33% of the districts across all 99 countries had more than 50% of their unvaccinated children living more than 2 hours from a health facility by foot. At the same time, many zero-dose children overall lived within 30 minutes of a health facility under a motorised scenario, indicating that proximity alone does not ensure vaccine uptake and that additional non-geographic barriers contribute to persistent zero-dose status. We provide a foundation for targeted vaccination strategies and interventions to bridge vaccination gaps and ensure equitable access to essential healthcare services.

Assessing walking and motorised travel times highlight where geographic barriers contribute to low vaccine uptake, while also pointing to settings where other factors play a larger role. These findings show where tailored interventions such as outreach services, mobile clinics or new facility placement may be required for locations that are difficult to reach. They also highlight the need to consider local contexts where conflict, insecurity or weak health system performance limit the availability or quality of services, even when geographic access is not a major barrier. Addressing both geographic and nongeographic barriers will help ensure that the children most in need are reached, reducing vaccination gaps and improving equitable access to essential healthcare.

## INTRODUCTION

Globally, there has been great success in delivering childhood immunizations, preventing millions of deaths each year from diseases such as diphtheria, tetanus and pertussis (DTP). However, progress has slowed in recent years [1] and even reversed since 2020, due to the COVID-19 pandemic and its associated disruptions. In 2023, an estimated 21 million children were un- or under-vaccinated [2].

This marks a slight decrease from 2021, when the figure reached 25 million, the highest number since 2009 [3]. Among these children, 18.1 million did not receive the first dose of the DTP vaccine (DTP1), indicating that many parts of the world still lack access to immunization and essential health care services [3].

To address current disparities in childhood immunization, the global Immunization Agenda 2030 (IA2030), seeks to maximise the impact of lifesaving vaccines by ensuring that everyone, everywhere should fully benefit from immunization [4]. By 2030, the Agenda aims to achieve a 90% coverage of essential childhood and adolescent vaccines and to reduce the number of children that do not receive any vaccines by half [5]. Additionally, GAVI, the Vaccine Alliance’s current strategy 5.0 (2021-2025), driven by the vision of leaving no one behind, aligned with the Sustainable Development Goals (SDGs) and IA2030, focuses on reaching those most marginalised and promoting the equitable and sustainable use of vaccines at the subnational level.

Despite developments in global policy and strategies, many studies that have looked at inequalities in vaccination coverage have focused on national level statistics [6–8]. Subnational estimates on the numbers of un- and under-vaccinated children are still limited [9,10]. Previous work has revealed significant inequalities at the district level [10–12] and finer spatial resolutions [13,14], but global comparative studies at the subnational level remain particularly rare. Studies that do assess the characteristics of un- and under-vaccinated children, geographical or otherwise, tend to focus on individual countries [15,16], or globally at the national level [17–19]. However, recent efforts have begun to characterise the number and distribution of zero-dose and under-immunised children across remote-rural, urban, and conflict-affected settings in low and middle-income countries (LMICs) [20].

Geographic access plays an important role in delivering equitable, essential healthcare services, including vaccinations, particularly in LMICs where infrastructure and transportation barriers often limit healthcare reach[17,21–24]. Studies across diverse settings have shown that travel time to health facilities directly impacts health outcomes. Examples of these are found in Ethiopia[25], Burkina Faso[26], Kenya [27], Uganda [28] and Nigeria[29], all of which found a link between greater distance to healthcare facilities and poorer health outcomes or reduced vaccination rates. These findings emphasise the critical need to reduce geographical barriers to achieve equitable access to healthcare. Broader analyses, such as that by Hierink et al. [30] and Ouma et al. [31], extend these insights by demonstrating how travel time affects timely access to surgical services across sub-Saharan Africa. Similarly, studies using geospatial data and modelling, such as Weiss et al. [32], highlight disparities in healthcare accessibility globally, showing that populations living further from health facilities are systematically underserved.

Understanding geographic access to health facilities for routine immunizations can help optimize service delivery for vaccine-preventable diseases like diphtheria, pertussis, tetanus, measles, polio, tuberculosis, and smallpox. Identifying underserved areas supports targeted vaccination efforts, efficient resource allocation, and informed policy decisions to close service gaps and achieve universal coverage. It also enables regional and global strategies to adapt as required to target limited resources effectively. However, there is limited work to date that characterises inequalities in such access to health services and the numbers and distribution of un- and under-immunised children both sub-nationally and on the global scale [27]. Such estimates can provide insights on global and regional trends, helping to address cross-border health risks, preventing the spread of diseases across countries and promote global equity in achieving immunization goals, while examination at the subnational level can identify geographic gaps in coverage, normally obscured by national averages [33].

Here, we explore the numbers and distribution of zero-dose children, defined as those not receiving the initial dose of the DTP1 vaccine [5], and geographic access to health care services, to assess spatial inequalities in immunization. We explore this both nationally and sub-nationally, to characterise differences in both walking and motorised travel-time across 99 LMICs, to provide consistent and comparable estimates between and within countries, that can be used to support global and regional decision making and vaccination campaigns.

## Data and methods

### Mapping children under the age of one and children unvaccinated for DTP1

To estimate the number of children under the age of one year old, we use the age and sex structured population counts available at 1x1km from the WorldPop database for 2021, for the 0-to-12-month age group [34]. These datasets have been adjusted to match the UN national total estimates [35] and spatially constrained to building footprints available for all the countries in the study area, following the findings and recommendations of Hierink et al. [36]. Details on the methods used to produce these are available elsewhere [37–39]. To estimate the numbers of unvaccinated children under the age of one, we used the methods described in Wigley et al. [20]. DTP1 vaccination coverage data from the Institute for Health Metrics and Evaluation (IHME) were aligned to the population data. To estimate the numbers of unvaccinated children at the grid square level the vaccination coverage was subtracted from one to calculate the rate of non-coverage, and then multiplied by the estimated population under 1 year of age.

These data were summarised at both the national and the second sub-national administrative unit level, which typically represents the district or county, referred to as districts in this paper, using adapted World Health Organization (WHO) health boundaries provided by GAVI in 2020. The main adaptation involved aligning and harmonising these boundaries to the population data at 1x1km. This means that all areas with data in the population dataset outside boundaries designated in the WHO dataset were allocated to district boundaries using nearest neighbour statistics. This created country and district boundaries that combined the maximum extents of the population data and WHO boundaries. WHO boundaries are commonly used by organisations such as GAVI to target their vaccination strategies and campaigns. Using these district boundaries supports the identification of subnational variations that otherwise are disguised at the national level. The resulting datasets were summarised for the 99 LMICs that had subnational boundaries at the district level and DTP1 vaccination coverage data available.

### Mapping children unvaccinated for DTP1 by travel-time to the nearest health facility

To estimate numbers of unvaccinated children by travel-time to the nearest health facility, we use datasets described in Weiss et al. [32]. These datasets are global gridded estimates of travel time to the closest health facilities, including hospitals and clinics from the public and private sector, at 1x1km resolution, using walking and motorised travel scenarios. In brief, these are calculated using a combination of travel speeds (minutes per metre) for each 1x1km grid cell based on the fastest travel mode intersecting the grid cell and using the geographical location of health facilities of any type from the Global Healthsites Mapping Project (https://www.healthsites.io/). The walking scenario assumes that no motorised transportation is available and therefore the travel time speed along all roads is limited to the walking speed of 5km per hour and crossing water without motorised transport with a travel speed of 1km per hour. The motorised scenario assumes a mixture of walking speeds off road and the use of motorised transportation, using a combination of road networks, railways, waterways, land cover types and topographical features, with varying speeds limits according to the type and the country boundaries.

Aligning to methodologies outlined in Wigley et al. [20] and Weiss et al. [32], we used travel-time categorisation to assess the distribution of children and un-vaccination coverage across various time intervals. Global benchmarks suggest that access to emergency health services should be between 30 minutes [40] to 1 hour [41] , while UNICEF [42], WHO [43] and the 2015 Lancet Commission on Global Surgery [44] recommend that access to health services should be within 2 to 3 hours. Therefore, a separate travel-time map was created for each band: 0 to 30, 30 to 60, 60 to 120, 120 to 180, and more than 180 minutes, for both walking and motorised transport. Cells within each of these ranges were assigned a value of 1. Cells in each band were then multiplied by cells recording the population under 1 year of age and un-vaccinated to estimate the number of unvaccinated children within each travel time band. Totals were then summarised by district using WHO health boundaries. The mean travel-time was used as a measure of central tendency in settled areas for each district, calculated using R version 4.3.2 [45]. Once these datasets were summarised, we compared them with vaccination coverage targets from the IA2030 [4] and benchmark travel times to identify suboptimal vaccination coverage and accessibility to services [27,43].

## RESULTS

### Geographic access to health facilities by district

Results show disparities in travel time to health facilities across the 18,424 districts in the 99 countries. The overall average travel time within the districts was 221 minutes under the walking travel time scenario and 44 minutes under motorised. In the walking scenario, 11.8% of districts had an average travel time between 0 and 30 minutes, 17.4% had an average between 30-60 minutes, and 46.1% had an average time exceeding 120 minutes, with one district in Papua New Guinea having an average of up to 8.1 days to reach a health facility. Additionally, all of the districts for 7 countries (Belize, Equatorial Guinea, Eswatini, Guyana, Lesotho, Namibia and Tajikistan) had an average travel time to health facilities of over 60 minutes, with Namibia and Tajikistan having over 70% of their districts showing an estimated average travel time of more than 180 minutes. In the motorised travel time scenario, 65.7% of districts had an average travel time between 0 and 30 minutes to a health facility, 18.1% had an average between 30 and 60 minutes, while 6.8% of districts had travel times exceeding 120 minutes. seen in **Figure S1**.

### Unvaccinated children for DTP1

In 2021 there were a total of 15.7 million unvaccinated children across the 99 countries analysed, representing 16 % of the total population of children under the age of one in these countries. India (2.9 million), Nigeria (2.3 million) and Indonesia (1.1 million) had the highest national totals. Proportionally, Somalia (58%) and Myanmar (52%) had the highest overall percentage of unvaccinated children, followed by Papua New Guinea (49%) and Guinea (42%).

Globally, only 40.3% of the districts studied were estimated to have achieved ≥90% vaccination coverage in 2021. Five countries had districts where over 70% of children were unvaccinated: Somalia (7.7%), Democratic Republic of the Congo (DRC) (1.5%), Nigeria (1.7%), Myanmar (1.2%), and the Philippines (1%). **Figure 1** shows the subnational disparities in DTP1 coverage in 2021.

Countries like the DRC, Ethiopia, Myanmar, Nigeria, Papua New Guinea, Philippines and Somalia, exemplify these contrasts, containing both high vaccination coverage districts, while also having some districts with the largest percentage of unvaccinated children among all the districts studied.

**Figure 1:**
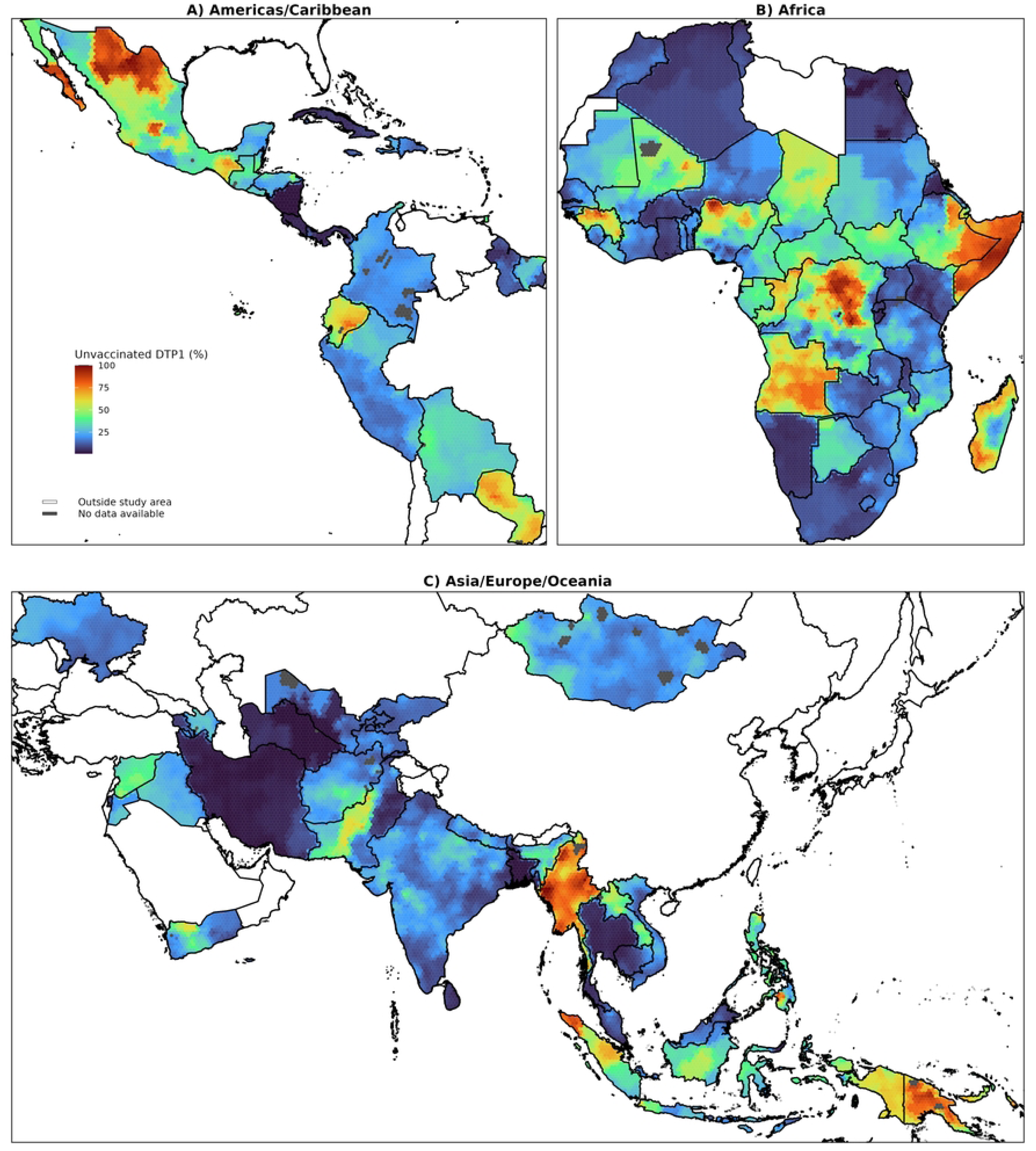
Spatial distribution of the estimated proportion of children under 1 year of age who did not receive the first dose of the DTP vaccine in 2021 by district, for Latin America and the Caribbean (A), Africa (B), and Asia, Europe, and Oceania (C). Data are visualized using mean values of hexagonal bins of 50 km. National boundaries: GADM v 4.1 (Retrieved from https://gadm.org)

### Unvaccinated children and geographic access to health facilities by district

We observed substantial disparities in the geographic distribution and travel time to health facilities of unvaccinated children, both across and within countries (**Table S1**). Using district level estimates of unvaccinated children under 1 year of age for DTP1 for 2021 and the mean travel time of each district by region (**Figures S2 and S3**), we found that the percentage of unvaccinated children generally increased as the mean travel time increased. There was substantial variation within the travel time bands for both walking and motorised travel scenarios in Africa and Asia. The presence of multiple outliers in the upper whiskers of the plots indicates that several districts had higher percentages of unvaccinated children. Although the effect of travel time was less pronounced in Latin America/Caribbean, this region had the largest number of districts (42%) with a mean travel time of ≥180 minutes.

Under the walking scenario, 48 districts across five countries: DRC (5,951 unvaccinated children), Nigeria (182,978), Somalia (31,917), Myanmar (21,478), and Indonesia (4,872), had mean travel times of 0-30 minutes with ≥50% of children unvaccinated. When considering the motorised scenario, the number of such districts increased markedly to 214 across Angola (5,668), DRC (30,564), Guinea (45,192), Indonesia (38,132), Myanmar (154,261), Nigeria (507,696), Papua New Guinea (24,962), the Philippines (22,381), and Somalia (129,955). Conversely, countries with districts having mean walking travel times ≥180 minutes and ≥50% unvaccinated children were Angola (63,219), Congo (15,816), DRC (78,327), Ethiopia (148,974), Madagascar (2,164), Mali (510), Myanmar (229,979), Papua New Guinea (63,705), and Somalia (200,862). For motorised travel times ≥180 minutes, these districts were located in Angola (14,258), DRC (13,370), Ethiopia (10,936), Myanmar (15,767), and Papua New Guinea (30,755).

Figure 2 further disaggregates these patterns by showing the distribution of district level mean travel times by country and region, alongside national DTP1 unvaccinated percentages indicated by the colour gradient. Across the regions, countries with higher district level mean travel times also tended to have higher percentage of unvaccinated children, as reflected in the colour gradient of the boxplots. Under the walking scenario, only 17 countries had the majority of their districts ≤60 minutes travel time to health facilities, yet some of these also had a large proportion of unvaccinated children e.g. Nigeria (30.6%) and Trinidad and Tobago (19%). In contrast, many countries with multiple districts experiencing the longest travel times, including Somalia (58%), Myanmar (52%) and Paraguay (26%), also had high national percentages of unvaccinated children.

**FIGURE 2.**
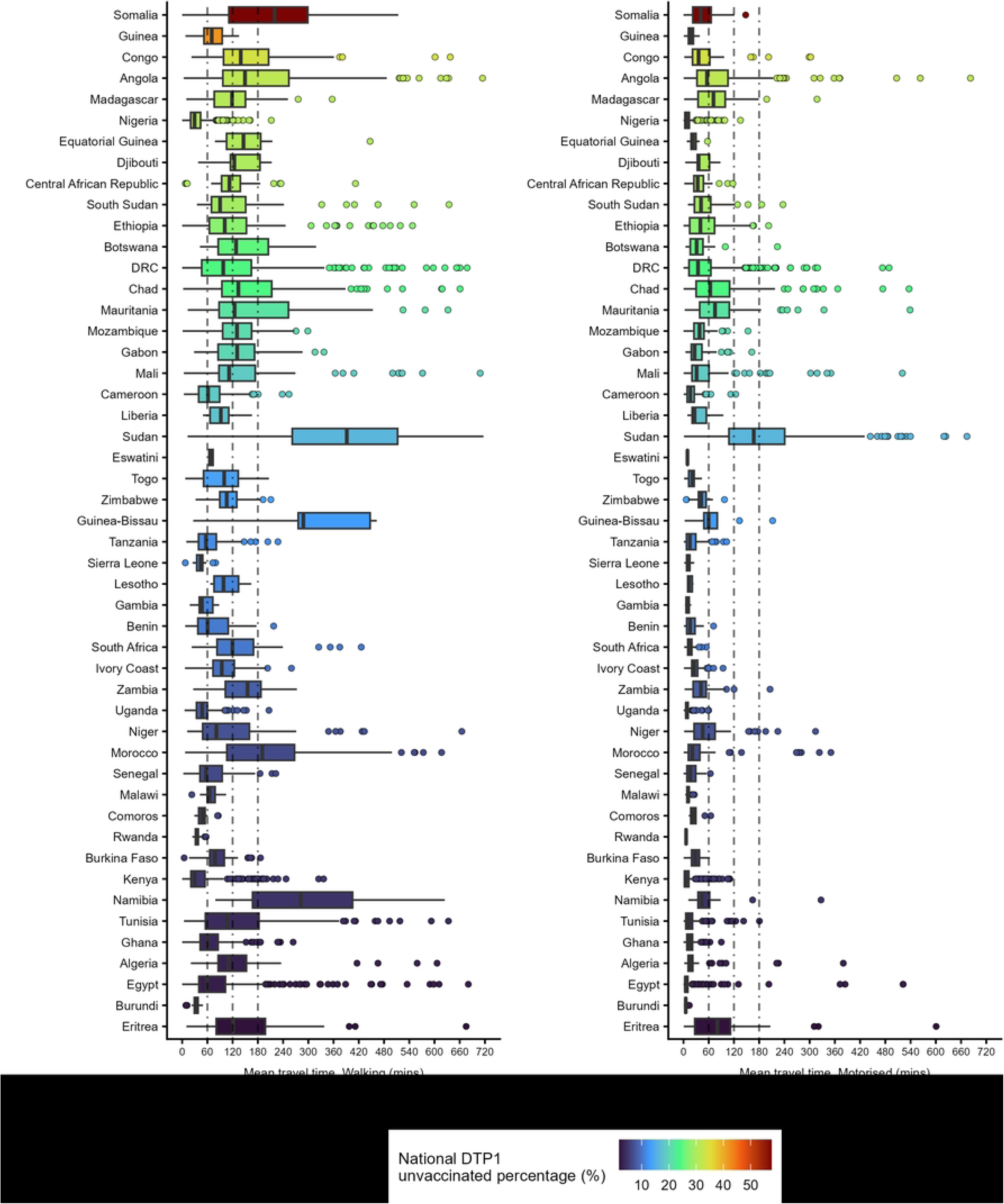
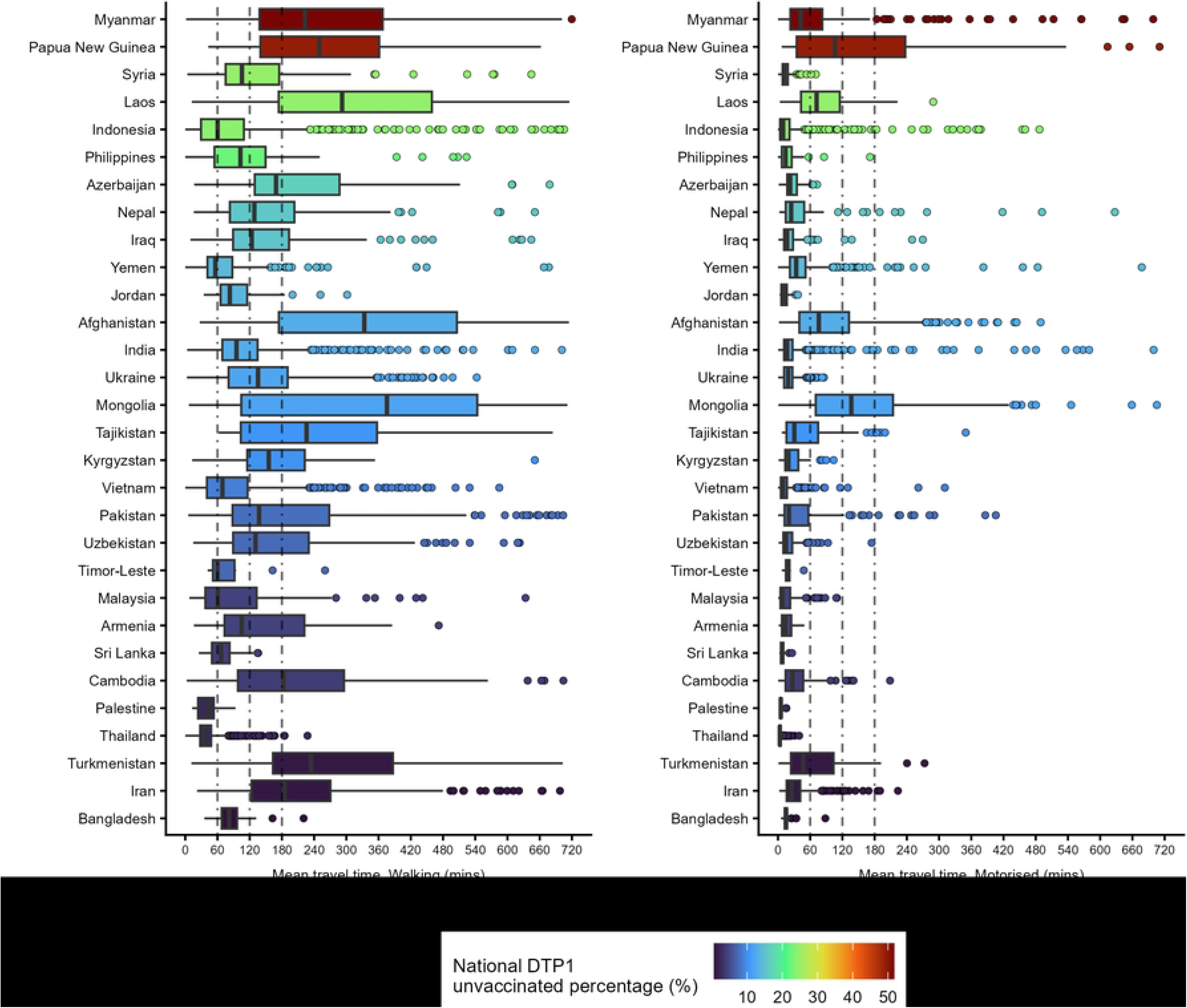
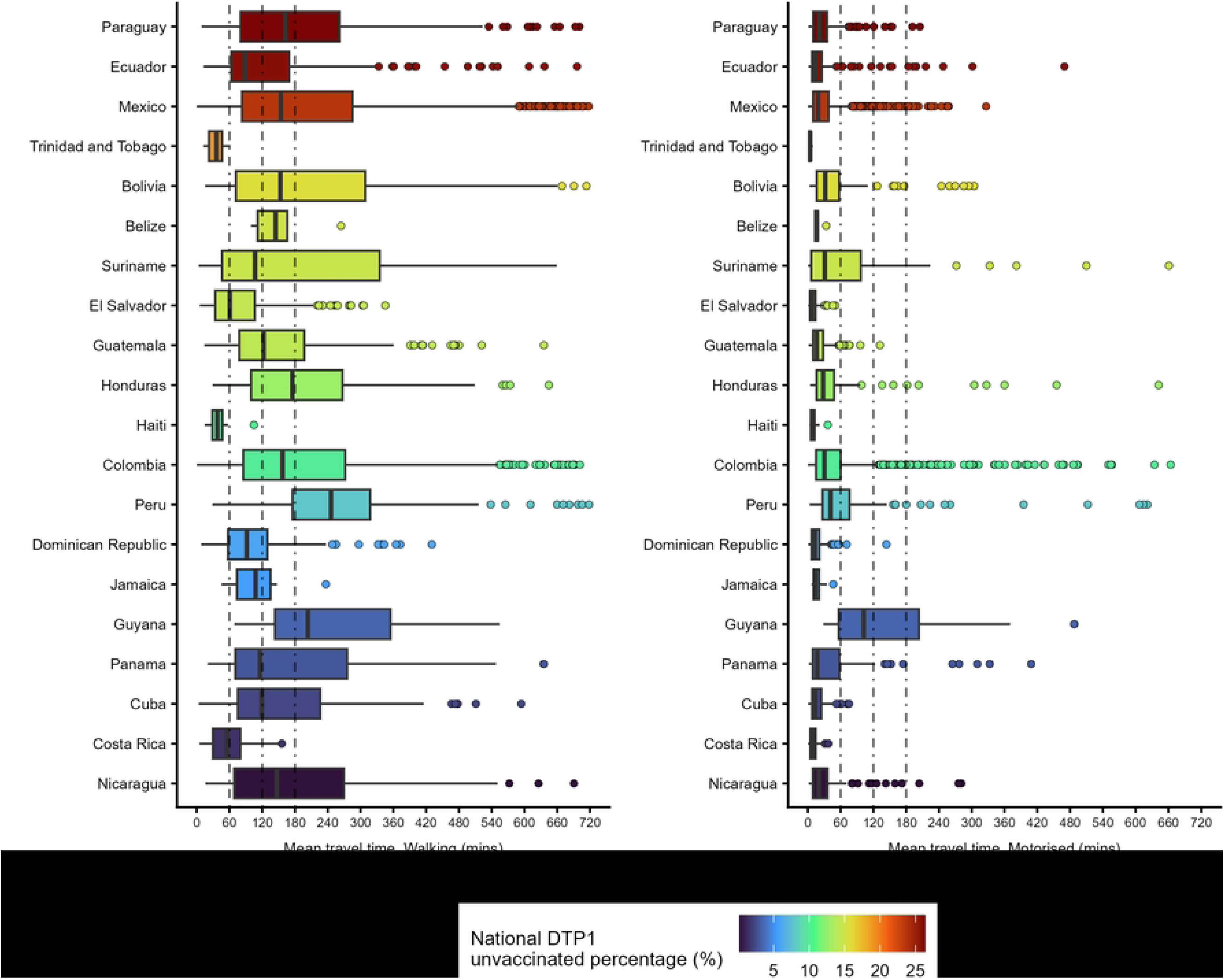
Box plots, showing the distribution of the mean travel-time for each district for each country by region Africa (2A), Asia/Europe/Oceania (2B) and Latin America/Caribbean (2C). The plot is limited to a maximum travel time of 720 mins. Additionally, the colour fill represents the estimated proportion of unvaccinated children in 2021 at the national level from blue (low) to red (high). Dots represent outlier units with values beyond the interquartile range. The dashed line shows the benchmark 1 hour (60 minutes), 2 hours (120 minutes) and 3 hours (180+ minutes) travel time to health facilities.

Comparing the spatial relationship between the district level percentage of children under one year old who were unvaccinated for DTP1 in 2021 and the mean travel time to health facilities under motorised and walking scenarios reveals clear contrasts across districts (Figure 3). These contrasts are particularly evident between districts with both low proportions of unvaccinated children and low mean travel times (shown in light yellow), and those with both high proportions of unvaccinated children and high mean travel times (shown in green). In many countries, however, there are districts where the proportion of unvaccinated children remains high despite relatively short mean travel times, as observed in Somalia, northern Nigeria, and central districts of the DRC.

**FIGURE 3.**
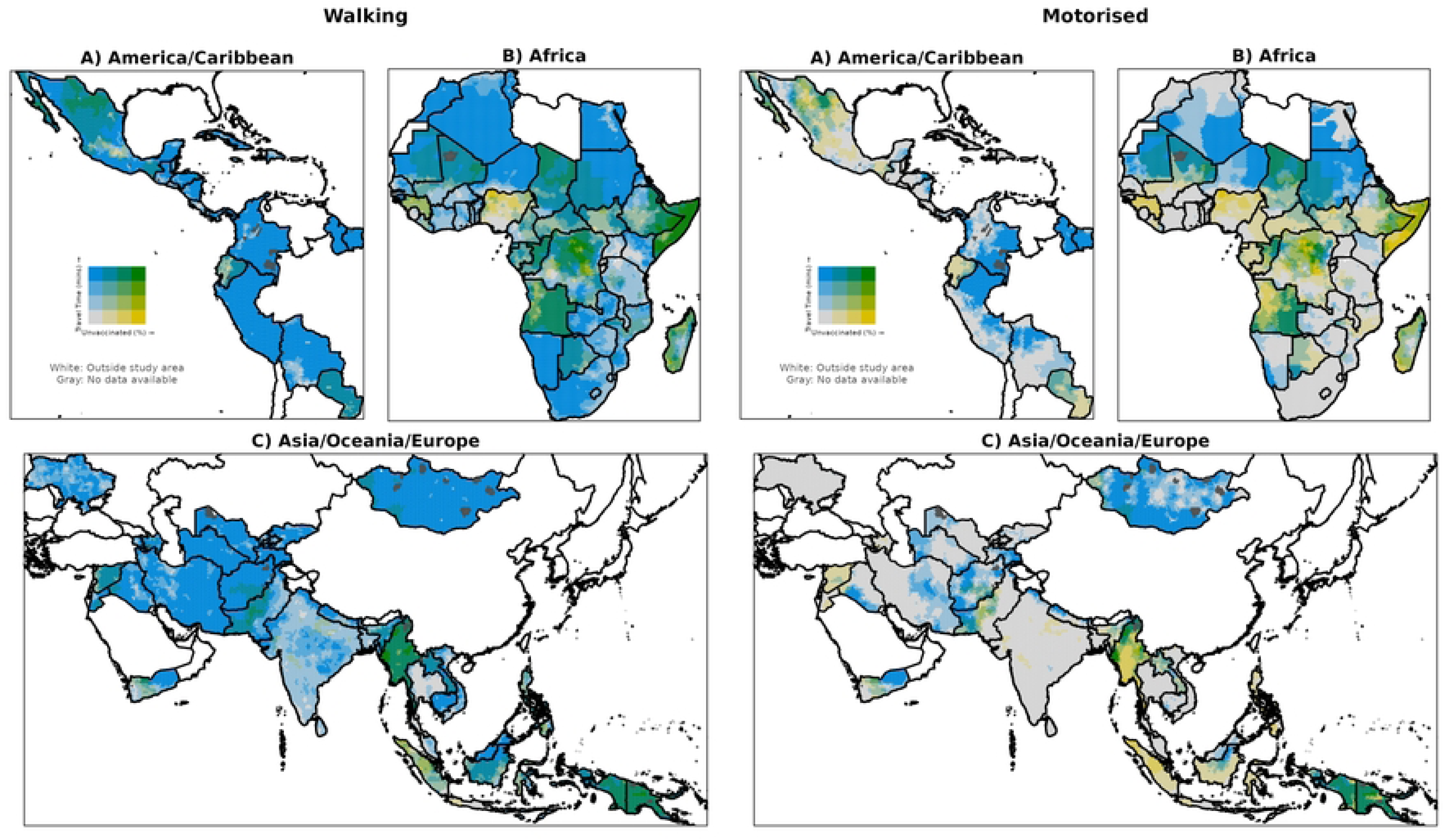
Bivariate map, showing high/low areas of the percentage of children under the age of one unvaccinated for DTP1 in 2021 and the mean travel-time for each district for A. Latin America/Caribbean, B. Africa, and C. Asia/Europe/Oceania. High/low categories are defined using break points of 60, 120, 180 and >180 mins travel time and 20, 40, 60 and >60% unvaccinated (DTP1). These are presented for the motorised (left panel) and walking (right panel) scenarios. Data are visualized using mean values of hexagonal bins of 50 km. National boundaries: GADM v 4.1 (Retrieved from https://gadm.org).

Figures 2 and **3** present summaries of district and national level patterns of unvaccinated children and mean travel time to health facilities, describing travel conditions within and across countries. Figures 4 and **5** provide a different perspective by examining the spatial distribution of unvaccinated children across travel time bands within districts. Figure 4 shows the proportion of unvaccinated children in each travel time band (0-30, 30-60, 60-120, 120-180, and ≥180 minutes), highlighting additional distinct spatial patterns of unvaccinated children across the study regions under both the walking and motorised scenarios. Presenting both walking and motorised scenarios allows the results to be interpreted in each specific national and sub-national context, as the relevance of each scenario depends on the transportation options realistically available in different settings.

Overall, under the walking scenario, 44% of unvaccinated children were located within 30 minutes of a health facility, meaning that 56% were farther than 30 minutes away, with at least 22% located ≥120 minutes from a facility. Under the motorised scenario, 83% of unvaccinated children were within 30 minutes of a health facility, although 4% were still found beyond 120 minutes. Breaking this down further by region, Figure 5 shows the proportion of unvaccinated children in each travel time band by country and region, highlighting contrasts both between and within countries under motorised and walking scenarios.

Under walking conditions, fewer than 43% of countries had a majority of their unvaccinated children within 30 minutes of a health facility. Furthermore, 39% of unvaccinated children lived more than one hour away from a health facility on foot, with Afghanistan, Papua New Guinea, Sudan, and Cambodia having the largest proportions (84%, 83%, 81%, and 78%, respectively). Seven countries, Guinea-Bissau, Sudan, and Afghanistan in Africa, and Cambodia, Laos, Yemen, and Papua New Guinea in Asia/Oceania, had more than 50% of their unvaccinated children living more than 180 minutes from a health facility on foot. Ony a few countries had a majority of their children living more than 30 minutes away from a health facility by motorized transport. These included Chad, Eritrea, Guinea Bissau, Madagascar and Sudan in Africa, and Afghanistan, Laos, and Papua New Guinea in Asia/Oceania. Overall, 9% of unvaccinated children lived further than one hour away by motorised transport, with Sudan, Afghanistan, Papua New Guinea, and Eritrea having the highest proportions (59%, 48%, 47%, and 42%, respectively).

**Figure 4.**
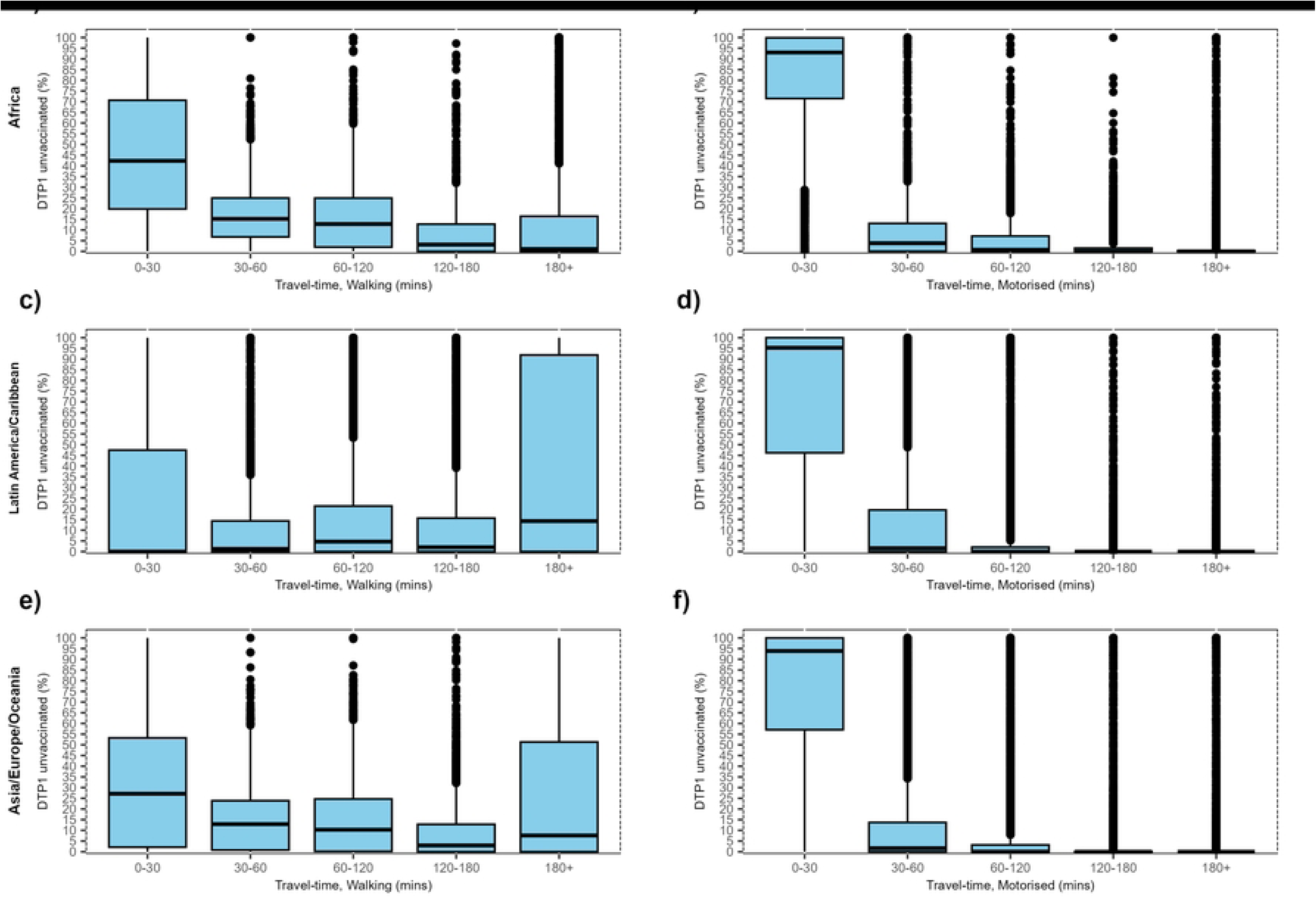
Box plots showing the percentage of children under 1 year of age unvaccinated for DTP1 grouped by the travel-time to health facilities for each district under the walking (a, c and e) and motorised scenario (b, d, and f) for Africa (a and b), Latin America/Caribbean (c and d) and Asia/Europe/Oceania (e and f).

**FIGURE 5.**
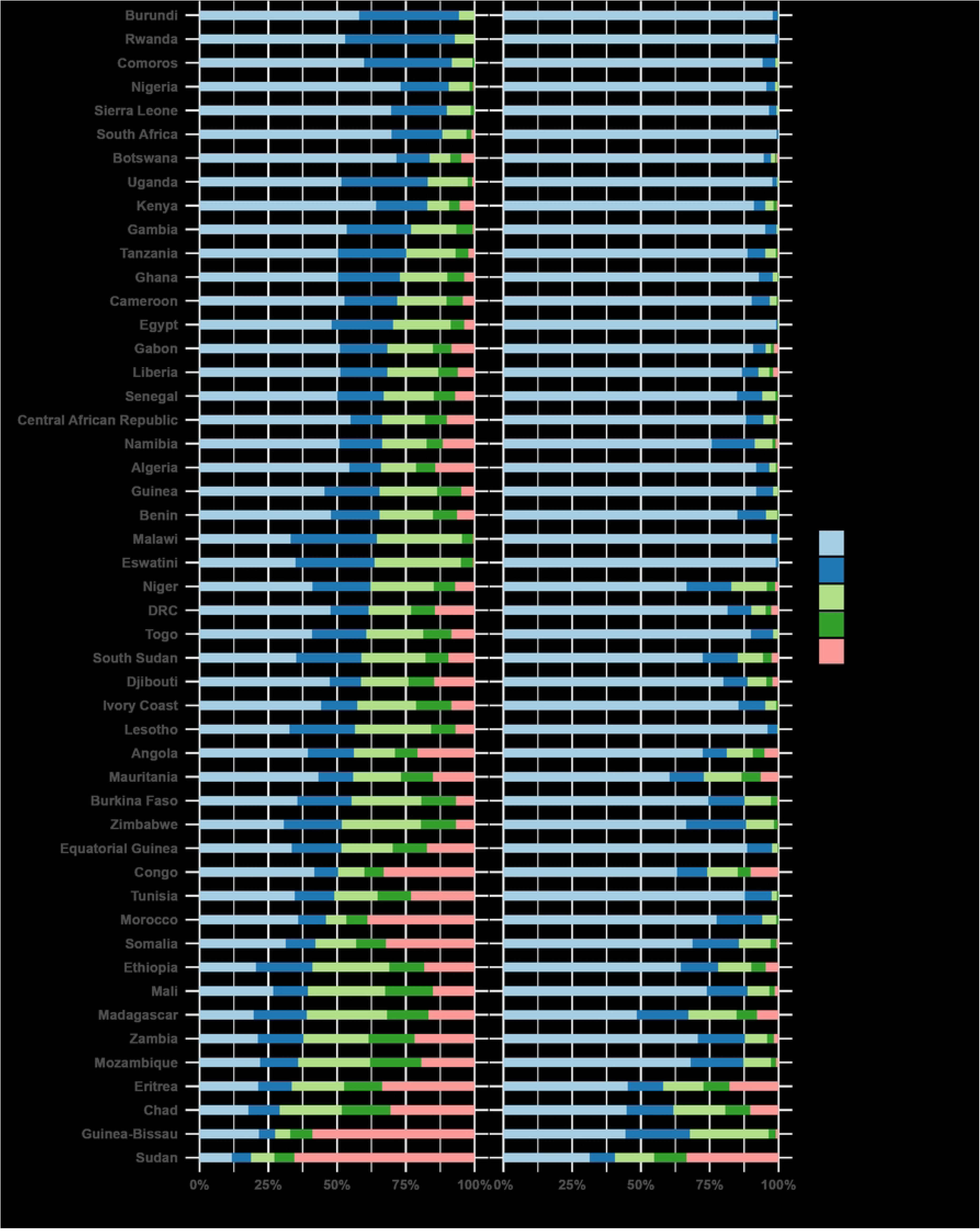
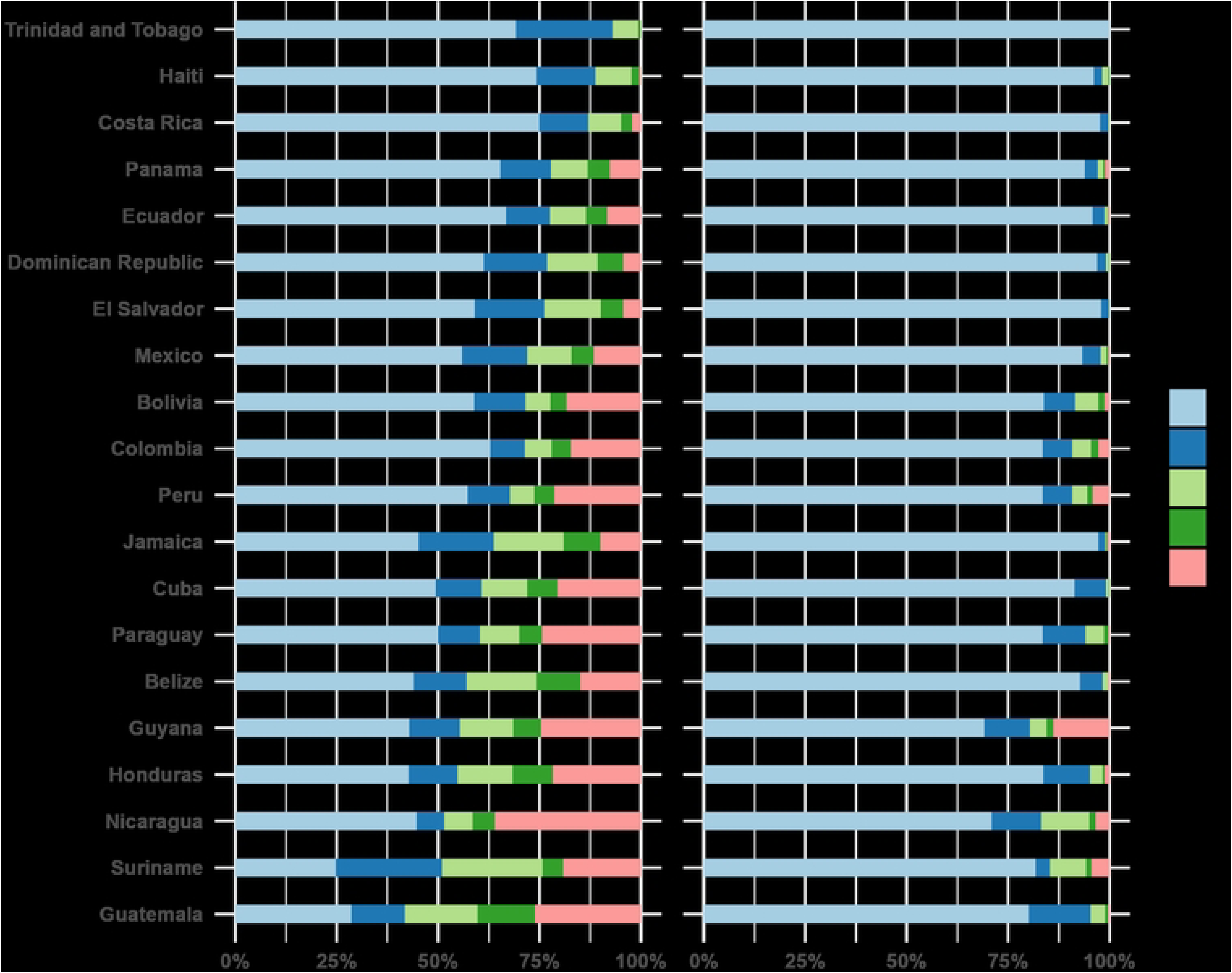
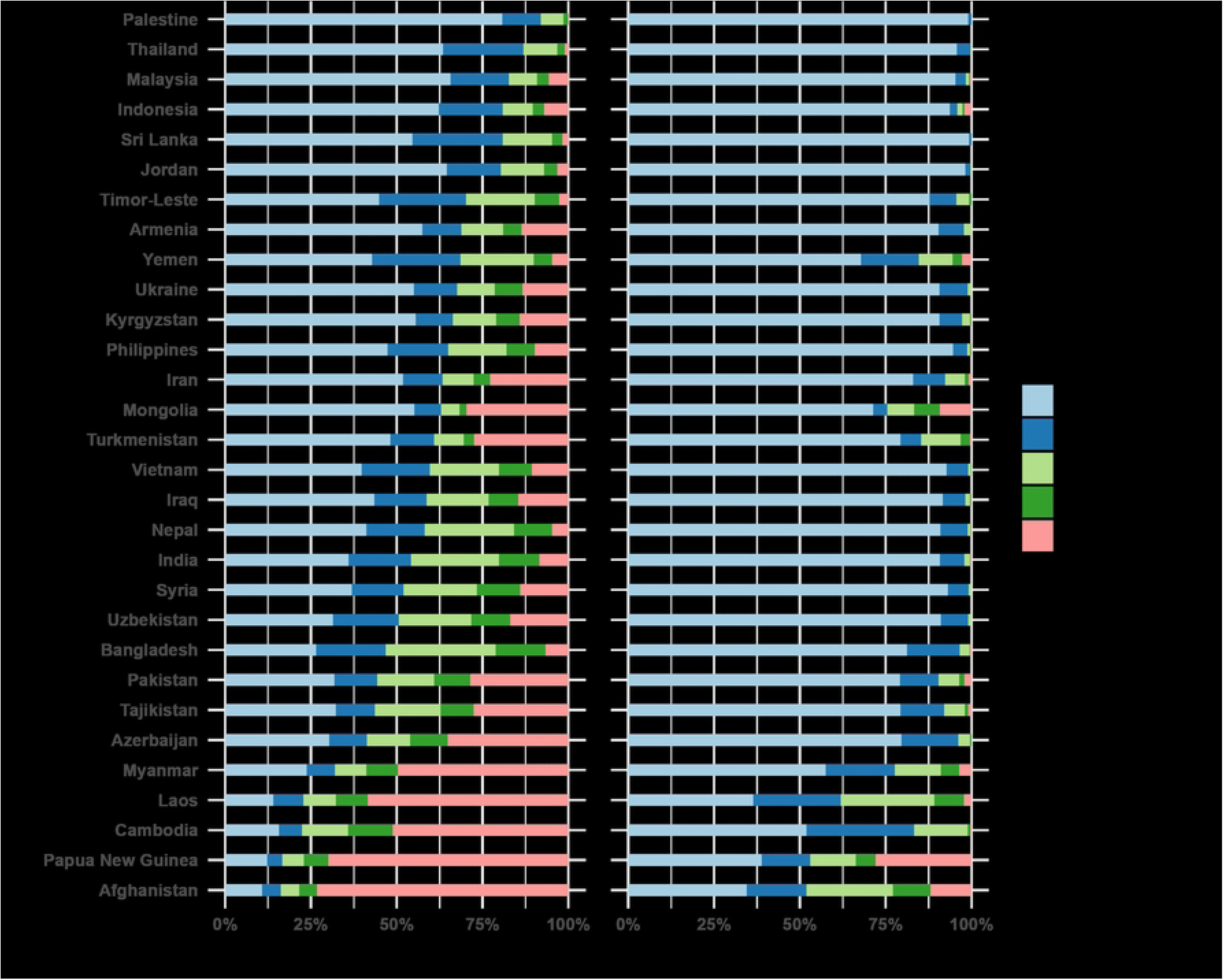
Stacked-bar charts, showing the percentage of children under the age of one unvaccinated for DTP1, split by travel-time bands, by region for all areas (A-C) under the walking (left) and motorised (right) scenario.

## DISCUSSION

To achieve a substantial reduction in the number of zero-dose children by reaching high and equitable coverage levels [4], immunization programs must become more effective at identifying underserved children. The results of this study highlight the variability in geographic access and unequal vaccination coverage of DTP1 among children under 1 year of age. The study shows that DTP1 vaccination efforts in 2021, a year affected by the COVID-19 pandemic [46], had a larger proportion of children unvaccinated, with noticeable subnational inequalities within and among countries. By incorporating small area measures of geographic access to health facilities, this study provides insights on subnational disparities in travel time and immunization, revealing important barriers that are often masked by national or regional summaries and that must be addressed to meet global vaccination goals.

We estimated that there were at least 15.7 million unvaccinated children in 2021, with disparities at both national and district levels. While some areas had high vaccination coverage, others had large proportions of unvaccinated children. Districts with larger populations tended to have higher absolute numbers of unvaccinated children despite coverage levels closer to national averages, whereas smaller, low-coverage districts often had fewer unvaccinated children, suggesting that population size may partly explain these patterns. Furthermore, substantial disparities in geographic access to health facilities across the 18,424 districts in 99 countries were observed. Under a motorised travel scenario, most districts had an estimated average travel time of 0-30 minutes, but under a walking scenario, nearly half of the districts faced over 120 minutes of travel time, with some reaching more than 8 days. A relationship between longer travel times and higher rates of unvaccinated children was observed in some countries (**Table 1S**), especially under walking scenarios, which further highlights the impacts of poor geographic access on immunization coverage[17,21,27,47,48]. Despite the availability of motorised transport options, affordability and accessibility barriers persist in many LMICs, where car ownership rates are low, many rely on informal or public transport, or walking to health facilities, with transport expenditures generally remain low [49,50]. Evidence suggests that walking often represents the dominant mode for accessing health facilities in certain settings[51], though the relative contribution of walking versus motorised transport can vary by context and facility type [28,52]

The implementation of effective routine immunization to meet established targets faces multiple challenges. One key issue is the suboptimal delivery of vaccines to remote areas, which is closely linked to low vaccination coverage in these locations [53,54]. This challenge can stem both from supply chain barriers that delay the physical movement of vaccines to remote facilities and from service delivery gaps that limit the ability of health workers to reach and immunise the target population[55]. This highlights the importance of addressing logistical and infrastructural barriers in remote regions. Hierink et al. [30] conducted a systematic scoping review and identified that limited geographical access to health facilities is also strongly associated with higher disease burdens in these areas. Multiple studies in specific countries and regions have identified that child immunization is significantly lower in areas that are further than 1 hour walking travel time from a facility, e.g. Niger [51], Kenya [27] and sub-Saharan countries [24]. Having consistent and comparable estimates between and within countries, supports global and regional decision-making processes. This also forms a basis for tracking progress towards reducing gaps and inequalities over time [56].

One limitation is the Modifiable Areal Unit Problem (MAUP), which arises when the results of the analysis are influenced by the spatial configuration and size of the geographic districts used. Our study used administrative boundaries that vary widely in shape and size, potentially affecting the outcomes of the spatial analyses [57,58]. Another limitation is the lack of detailed health facility information, including factors such as whether and which immunization services are provided.

Comprehensive data on the types of healthcare facilities and the specific services they provide would have allowed us to select only the most appropriate health facilities related to the immunization of children and provide better estimates, especially in conflict affected settings in LMICs[59]. Additionally, we did not consider vaccination outreach or mobile services by community health workers and those outside of formal facilities [60], which can have an impact in access to immunisation in rural and under-reached communities. The accuracy of population data is critical for spatial and accessibility analyses, yet our study faced uncertainties related to population estimates. Following the guidance in Hierink et al. [36], this work used population estimates where mapping was constrained to satellite imagery-defined settlements and building footprints. However, such satellite-derived datasets are not perfect [61], and this can mean some populations missed from analyses in some areas or erroneously mapped in others. Hierink et al. [36] explored the effect of population data on geographic access in sub-Saharan Africa and identified the large differences that exist in the different population datasets while assessing accessibility, especially in areas of sparse population.

Assumptions made in travel time modelling present another limitation. Travel times are based on estimates for 2019, which may have changed substantially in some areas by our study year of 2021 with the development of new roads or other transport infrastructure. Moreover, while our analysis primarily focused on geographical accessibility, it did not consider other critical dimensions of healthcare access, such as financial, cultural, and organizational factors [62]. Additionally, the interpretation of motorised access findings should consider the variability in access to vehicles and public transport options across different regions. Countries with low levels of car ownership are likely to rely more heavily on public transport or non-motorised modes of travel, such as cycling e.g. Dowhaniuk [63], increasing the travel time. Furthermore, the extent to which different modes of transport are used varies depending on multiple factors and often involves a mix of methods[52]; therefore, basing results solely on walking or motorised transport may overestimate or underestimate travel time .There are also uncertainties related to the vaccination coverage modelling as the data is estimated at a 5x5km from surveys undertaken across multiple years, and this may have added uncertainty and resulted in an over or under-estimation of coverage in some locations [12]. Lastly, our analysis was restricted to a single year, 2021, which may not adequately capture trends or changes over time. Healthcare access and demographic characteristics are dynamic, and significant changes may have occurred since the study period that could influence the relevance of our findings, especially as the year we worked on was impacted by the COVID-19 pandemic. Continuous data updates are essential to understand how healthcare access evolves and to ensure that policies and interventions remain effective over time.

Despite the inherent uncertainties, this analysis provides valuable insights into inequalities in access to vaccination, highlighting critical information needed by public health policymakers, government health officials, and global health organizations such as UNICEF, WHO and GAVI, working on immunisation programmes. The main patterns identified in this study can be viewed through two key typologies of districts. First, areas with low vaccination coverage but relatively short travel times, e.g. in DRC, Nigeria, Somalia, and Myanmar, which may reflect contexts where conflict, insecurity, or weak health system performance limit the availability or quality of services, even when geographic access is not a major barrier. Second, districts with both low coverage and long travel times likely correspond to rural and remote settings where physical accessibility, poor transport infrastructure, and limited health system capacity could constrain service delivery. Recognising these distinct contexts is important for tailoring interventions, to strengthen service delivery and outreach strategies in the former, while prioritising infrastructure, mobile clinics, and community-based delivery in the latter. Understanding regional variations in vaccination coverage enables stakeholders to develop targeted immunization strategies that address the specific needs of underserved and hard-to-reach populations. Overlaying modelled small area estimates of vaccination rates with health facility access data also gives health practitioners a good sense of where investment in health facilities is needed, or whether out of facility services, such as the use of Community Health Extension workers or mobile and temporary vaccination posts, may be required (as per the recommendations of Gibson et al. [60]). These findings can inform policies and programs at local, national, and international levels, guiding resource allocation and intervention planning to ensure equitable access to essential vaccinations and strengthen global immunization efforts. Furthermore, the data applicability extends beyond the district level use, as it can be adapted to different administrative levels, such as health districts, and integrated with other demographic and socioeconomic characteristics. By incorporating these additional factors, stakeholders can better address broader barriers to immunization, advancing progress towards universal vaccine coverage.

## Data Availability

1) Population data can be found here: https://doi.org/10.5258/SOTON/WP00743 2) Vaccination data can be found here: https://www.healthdata.org/research-analysis/health-topics/vaccine-coverage-data 3) Travel time data can be found here: https://malariaatlas.org/project-resources/accessibility-to-healthcare/ 4)National boundaries can be found here: https://gadm.org 5)Subnational administrative boundaries data, for legal/ownership reasons, we are not able to share.

## ETHICS DECLARATIONS

Ethics approval for this article was obtained from the Ethics Committee at the Faculty of Environmental and Life Sciences, University of Southampton, United Kingdom (Ethics number: 50248.A1). Consent to participate is not applicable.

## ACKNOWLEDGMENTS

The authors thanks to Carla Pezzulo for her supportive discussion on the topic and the WorldPop PMO team for their support. Additionally, the authors acknowledge the use of the IRIDIS High Performance Computing Facility, and associated support services at the University of Southampton, in the completion of this work.

